# COVID-19 diagnosis and study of serum SARS-CoV-2 specific IgA, IgM and IgG by chemiluminescence immunoanalysis

**DOI:** 10.1101/2020.04.17.20064907

**Authors:** Huan Ma, Weihong Zeng, Hongliang He, Dan Zhao, Yunru Yang, Dehua Jiang, Peigen Yingjie Qi, Weihuang He, Changcheng Zhao, Ruting Yi, Xiaofang Wang, Bo Wang, Yuanhong Yun Yang, Arnaud John Kombe Kombe, Chengchao Ding, Jiajia Xie, Yong Gao, Linzhao Cheng, Yajuan Li, Xiaoling Ma, Tengchuan Jin

## Abstract

**Background:** The pandemic of the Severe Acute Respiratory Syndrome Coronavirus 2 (SARS-CoV-2) is causing great loss. Detecting viral RNAs is standard approach for SARS-CoV-2 diagnosis with variable success. Currently, studies describing the serological diagnostic methods are emerging, while most of them just involve the detection of SARS-CoV-2-specific IgM and IgG by ELISA or “flow immunoassay” with limited accuracy.

**Methods:** Diagnostic approach depends on chemiluminescence immunoanalysis (CLIA) for detecting IgA, IgM and IgG specific to SARS-CoV-2 nucleocapsid protein (NP) and receptor-binding domain (RBD) was developed. The approach was tested with 216 sera from 87 COVID-19 patients and 483 sera from SARS-CoV-2 negative individuals. The diagnostic accuracy was evaluated by receiver operating characteristic (ROC) analysis. Concentration kinetics of RBD-specific serum antibodies were characterized. The relationship of serum RBD-specific antibodies and disease severity was analyzed.

**Results:** The diagnostic accuracy based on RBD outperformed those based on NP. Adding IgA to a conventional serological test containing IgM and IgG improves sensitivity of SARS-CoV-2 diagnosis at early stage. CLIA for detecting RBD-specific IgA, IgM and IgG showed diagnostic sensitivities of 98.6%, 96.8% and 96.8%, and specificities of 98.1%, 92.3% and 99.8%, respectively. Median concentration of IgA and IgM peaked during 16-20 days after illness onset at 8.84 μg/mL and 7.25 μg/mL, respectively, while IgG peaked during 21-25 days after illness onset at 16.47 μg/mL. Furthermore, the serum IgA level positively correlates with COVID-19 severity.

**Conclusion:** CLIA for detecting SARS-CoV-2 RBD-specific IgA, IgM and IgG in blood provides additional values for diagnosing and monitoring of COVID-19.

**Summary:** Chemiluminescence immunoanalysis of SARS-CoV-2 RBD-specific serum IgA as well as IgM and IgG improves accuracy of COVID-19 diagnosis. Concentration kinetics of serum RBD-specific IgA, IgM and IgG are revealed. Serum IgA levels positively correlate with COVID-19 severity.

## Introduction

At the end of 2019, a novel coronavirus (2019-nCoV or SARS-CoV-2) emerged in Wuhan, Hubei Province in China, causing a new type of coronavirus disease now named as COVID-19[1]. The virus spread globally and became a public health emergency and pandemic declared by the World Health Organization[2]. Among the seven coronaviruses known to cause human diseases, the severe acute respiratory syndrome (SARS) virus broke out in 2003[3] and Middle East Respiratory Syndrome (MERS) virus in 2012[4], COVID-19 is pathologically similar to but different from SARS and MERS is expected to cause great impact on human society since World War II[5]. Reliable and effective diagnosis of SARS-CoV-2 and treatment of COVID-19 are urgently needed.

Detection of SARS-CoV-2 viral RNA by methods such as RT-qPCR supplemented by chest CT imaging is the primary method for clinical diagnosis of COVID-19[6, 7]. However, the difficulty to obtain high-quality and consistent throat or nasal swab samples, and the low viral load at the late stage of infection, both challenges resulted in a sensitivity below 70%[8-12]. Therefore, there is an urgent need for more reliable and rapid diagnostic approach to screen SARS-CoV-2 infected people including those who do not have overt symptoms. A serological test of virus-induced antibody production has unique advantages in clinical diagnostics, especially for identifying people who acquired immunity against pathogens without noticeable symptoms[13]. When the virus invades host, the body produces large amounts of immunoglobulin (Ig) by the immune system and releases them into blood, including IgG, IgM and IgA[14]. It is known that IgM is normally the first antibody to be produced in response to the virus invasion[14]. IgG is a major class of immunoglobulins found in the blood, comprising 75% of total serum immunoglobulins and has long-term immunity and immunological memory[14, 15]. Therefore, measuring the viral antigen-specific of IgM and IgG in combination has been used in various serological tests for detecting SARS-CoV-2 infection as previously used for SARS and other coronaviruses[9, 10, 13, 16-20]. In contrast, IgA that is mainly produced in mucosal tissues to stop virus invasion and replication but also present in blood (∼15% of total immunoglobulins in blood)[21], has not been widely used in serological tests for detecting coronavirus infection. IgA kinetics and roles in anti-viral immunity are even less known. Currently, only a few published studies reported diagnosis of COVID-19 by using ELISA or “flow immunoassay” for detection of serum IgM and IgG with limited accuracy[9, 10, 16-19]. Although detection of SARS-CoV-2 specific IgA in serum was reported in recent papers or a preprint[11, 22, 23], The kinetics of antibody responses in COVID-19 remains undefined, specifically for IgA production.

In this investigation, SARS-CoV-2 specific IgA as well as IgM and IgG in 216 sera from 87 COVID-19 patients and 483 negative sera were evaluated using chemiluminescence immuno-analysis (CLIA), we demonstrated that the approach based on CLIA of SARS-CoV-2 RBD antibodies have improved diagnostic sensitivity and specificity. Kinetics of each antibody isotype and relationship of serum antibodies and disease severity were also revealed.

## Methods

### Patients and clinical samples

This study was approved by the Medical Ethical Committee of the First Affiliated Hospital of USTC and the First Affiliated Hospital of Anhui Medical University. Patient information is listed in **supplementary Table 1** with a detailed description. Patient classification was defined according to the New Coronavirus Pneumonia Prevention and Control Program (7th edition) published by the National Health Commission of China. This study enrolls a total of 87 cases of RT-qPCR confirmed COVID-19 patients, who were admitted to the First Affiliated Hospital of USTC Hospital or the First Affiliated Hospital of Anhui Medical University between Jan 26 and Mar 5, 2020. Their blood samples were collected during routine clinical testing. For all information of the enrolled patients were obtained from the clinical records.

**Table 1.** Comparisons of sensitivity, specificity and overall agreements of RBD-specific IgA, IgM, and IgG kit and their combinations for diagnosing SARS-CoV-2.

| Antibody type | Sensitivity |  | Specificity |  | Overall agreement |  |
| --- | --- | --- | --- | --- | --- | --- |
|  | % | n/total | % | n/total | % | n/total |
| IgA | 98.6 | 213/216 | 98.1 | 474/483 | 98.3 | 687/699 |
| IgM | 96.8 | 209/216 | 92.3 | 446/483 | 93.7 | 655/699 |
| IgG | 96.8 | 209/216 | 99.8 | 482/483 | 98.9 | 691/699 |
| IgA and IgM | 95.8 | 207/216 | 90.7 | 438/483 | 92.3 | 645/699 |
| IgA and IgG | 96.3 | 208/216 | 97.9 | 473/483 | 97.4 | 681/699 |
| IgM and IgG | 94.9 | 205/216 | 92.1 | 445/483 | 93.0 | 650/699 |
| IgA and IgM and IgG | 94.4 | 204/216 | 90.5 | 437/483 | 91.7 | 641/699 |
| IgA or IgM | 99.5 | 215/216 | 99.8 | 482/483 | 99.7 | 697/699 |
| IgA or IgG | 99.1 | 214/216 | 100 | 483/483 | 99.7 | 697/699 |
| IgM or IgG | 98.6 | 213/216 | 100 | 483/483 | 99.6 | 696/699 |
| IgA or IgM or IgG | 99.5 | 215/216 | 100 | 483/483 | 99.9 | 698/699 |

Sera as negative controls were collected in order to evaluate the diagnostic accuracy. This cohort contains 330 archived sera from healthy donors (samples collected before October 2019), 138 interfering sera from no-COVID-19 patients with different underlying diseases, and fifteen sera from once suspected cases (RT-qPCR negative but had typical manifestation of pneumonia). All sera were stored at −20°C before use.

### Molecular cloning, protein expression and purification

Briefly, the viral nucleocapsid protein (NP) was expressed using *E. coli* and purified with Nickel column and hydrophobic-interaction column. The SARS-CoV-2 RBD protein was expressed using HEK293 cell and purified from cell supernatant using Protein A column.

### Chemiluminescence immuno-analysis (CLIA) for SARS-CoV-2 diagnosis

Briefly, the purified NP or RBD viral antigens were coated onto magnetic particles for catching SARS-CoV-2 specific IgA, IgM and IgG in sera. A second antibody that recognizes IgA, IgM or IgG conjugated with acridinium (which can react with substrates to generate a strong chemiluminescence) was used for detecting the IgA, IgM or IgG caught by antigen, respectively. The detected chemiluminescent signal over background signal was calculated as relative light units (RLU). Such collection contains all contents for CLIA of antigen-specific immunoglobulin is called kit here. Serum samples were collected by centrifugation of whole blood in test tubes at room temperature for 15 min. Prior to testing, serum samples were treated with a final concentration of 1% TNBP and 1% Triton X-100 to completely denature any potential viruses[24]. Virus-inactivated sera were then diluted 40 times with dilution buffer and subjected to testing at room temperature. RLU was measured using a fully automatic chemical luminescent immunoanalyzer, Kaeser 1000 (Kangrun Biotech, Guangzhou, China).

### RBD-specific Antibody standards preparation

SARS-CoV-2 RBD was immobilized to agarose beads by using *CNBr-activated Sepharose™ 4B* reagent (GE Healthcare), then column filled with the RBD coupled agarose beads were employed to purify RBD-specific IgA, IgM and IgG antibodies from a serum pool of recovering patients (a manuscript in preparation). The concentrations of purified antibodies were determined using Bradford method (using bovine serum albumin protein as a standard). These antibodies were used to make a standard curve for each antibody detection to quantify the absolute antibody amounts in serum.

### Statistical analysis

Receiver operating characteristic (ROC) analysis was conducted using MedCalc software to determine the optimal cut-off value (criterion) and evaluate the diagnostic value of NP- or RBD-specific IgA, IgM and IgG detection. The specificity and sensitivity of the antibody detection were calculated according to the following formulas:

Specificity (%) = 100 × [True negative / (True Negative + False Positive)];

Sensitivity (%) = 100 × [True Positive / (True Positive + False Negative)];

Overall agreement (%) = (True negative + True Positive) / Total tests.

In order to analyze the correlation of serum antibody levels and age with disease severity, we first used the Kruskal Wallis test[25] to test if there is any significant difference of antibody levels and age among the three groups (Mild, Moderate, Severe). Then Dunn’s test[26] was used to perform a pair-wise test between each group, and Benjamini-Hochberg procedure[27] was used to adjust p-values. All the above analyses used R software version 3.6.1[28]. A p value less than 0.05 was judged statistically significant.

## Results

### SARS-CoV-2 RBD is better than NP for COVID-19 diagnosis by antibody detection

Highly purified SARS-CoV-2 NP and RBD proteins (**supplementary Figure 1**) were employed to make a set of CLIA kits (hereinafter referred to as “kit”) for detecting the presence of NP- and RBD-specific IgA, IgM and IgG, respectively. A cohort of 216 sera from 87 SARS-CoV-2 infected patients together with 20 interfering sera as negative controls were tested by both NP and RBD kits. ROC analysis was performed, the NP IgA, IgM and IgG kit showed diagnostic sensitivities of 89.8%, 78.2% and 95.8%, and specificities of 85.0%, 95.0% and 100% respectively (**Figure 1A-C**). However, the RBD IgA, IgM and IgG kit showed higher diagnostic sensitivities of 97.2%, 93.1% and 96.8%, and specificities of 100%, 90.0% and 100%, respectively (**Figure 1D-F**). We conclude that detection of RBD-specific antibodies provide a better diagnostic accuracy than that of NP-specific antibody.

**Figure 1.**
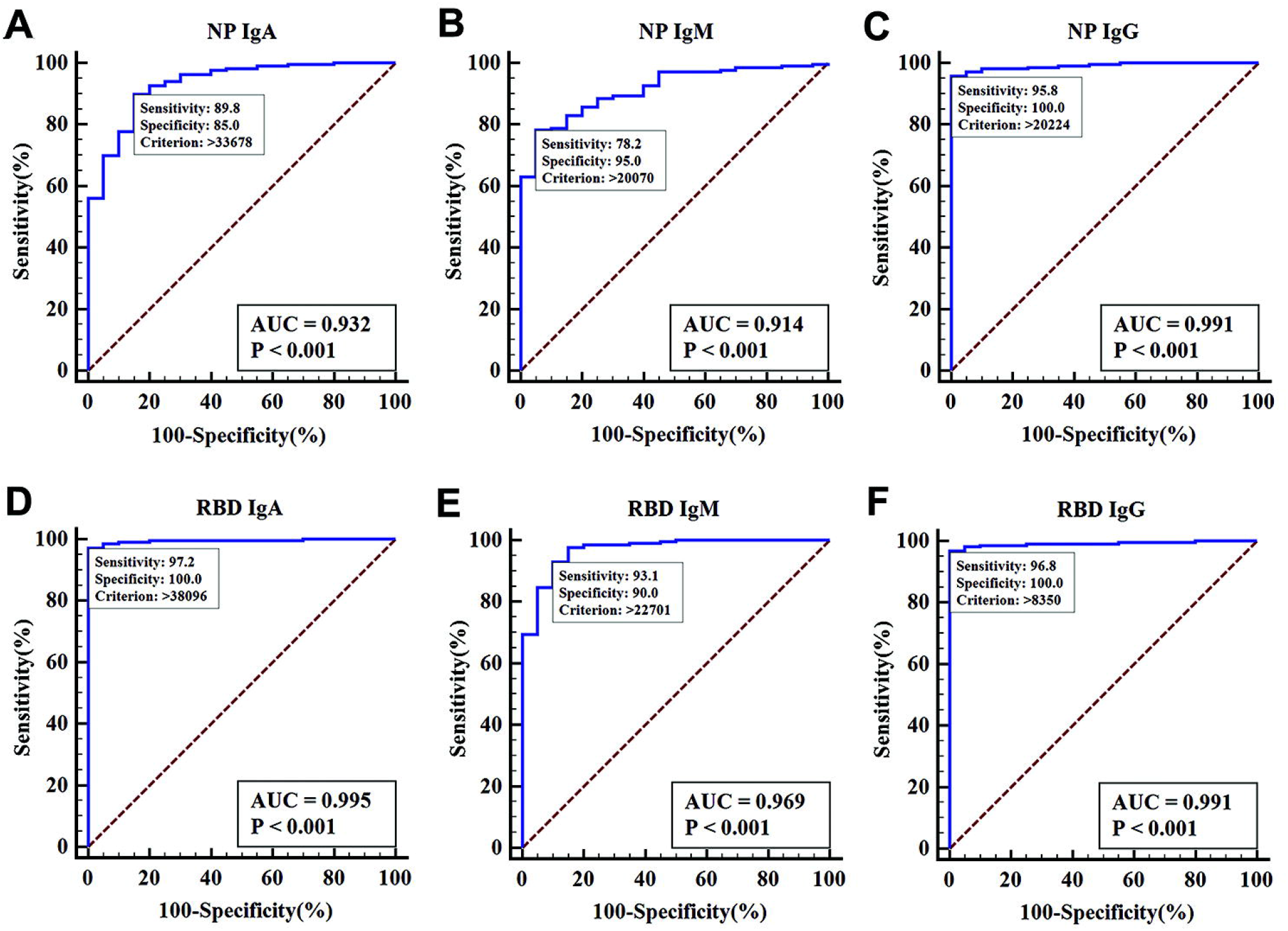
Comparison of NP- and RBD-based CLIA kits. The receiver operating characteristic (ROC) curve analysis for SARS-CoV-2 diagnosis by NP-specific IgA, IgM, and IgG kit (A, B, and C, respectively), and RBD-specific IgA, IgM, and IgG kit (D, E, and F), respectively. Twenty interfering sera and 216 sera from 87 SARS-CoV-2 infected patients were tested. AUC, area under the curve of ROC.

### Adding IgA to serological CLIA improves accuracy of SARS-CoV-2 diagnosis

To further evaluate the diagnostic accuracy of the RBD-specific IgA, IgM and IgG kit, a total of 483 sera including 330 healthy sera, 138 interfering sera and 15 sera from once suspected cases were tested as negative controls, respectively. Testing results were converted to scatter plots (**Figure 2A-C**), RBD-specific IgA, IgM and IgG CLIA kit showed diagnostic sensitivities of 98.6%, 96.8% and 96.8%, and specificities of 98.1%, 92.3% and 99.8%, respectively (**Figure 2D-F**). The sensitivities, specificities and overall agreements of the RBD-specific IgA, IgM or IgG kit and their combinations are also summarized (**Table 1**). When combining the of RBD IgA and IgG kit, the sensitivity, specificity and overall agreement elevated to 99.1%, 100% and 99.7%, respectively, which was much better than those when IgM and IgG kit were combined. IgA detection provides additional values for COVID-19 diagnosis.

**Figure 2.**
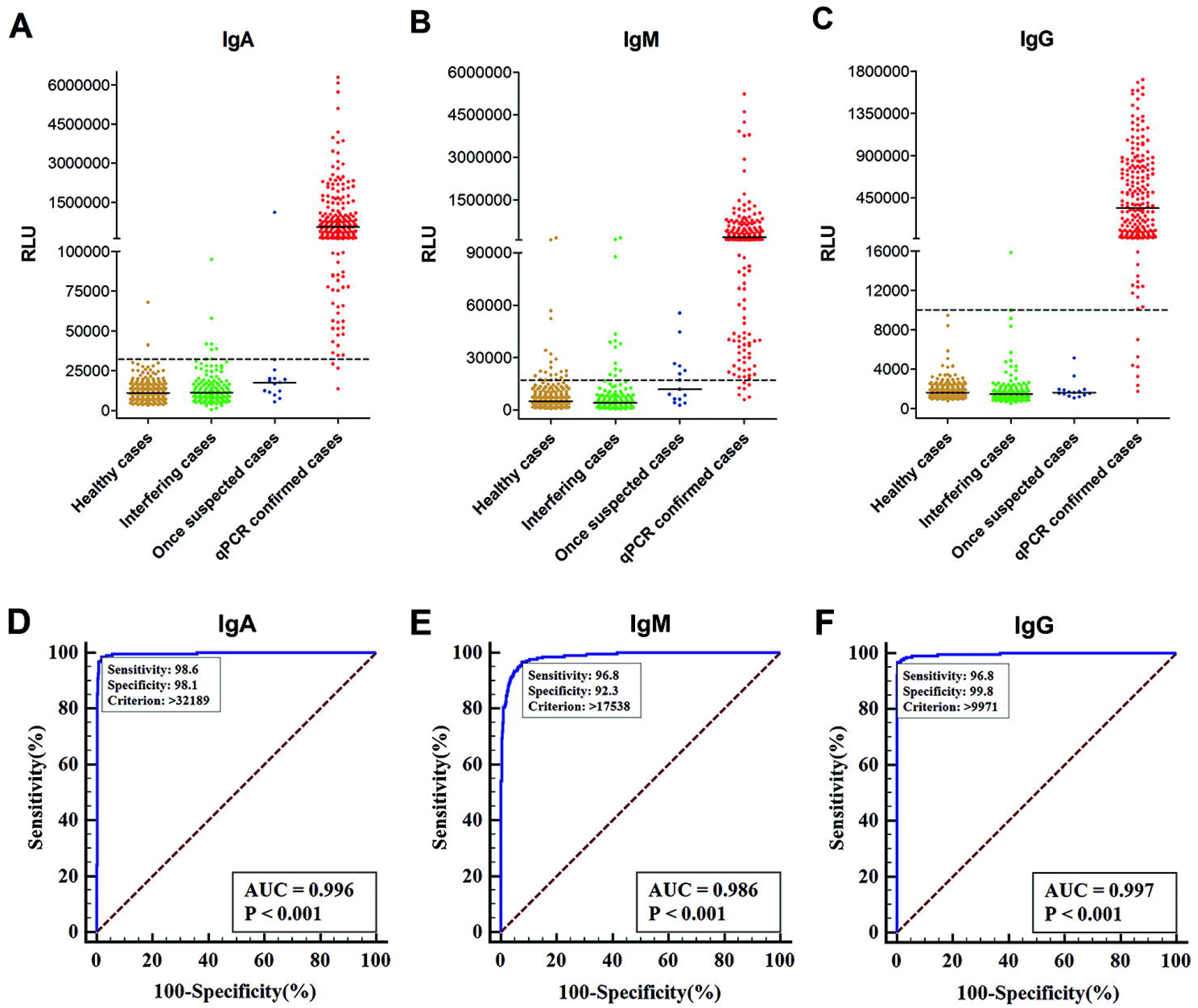
Testing results and diagnostic analysis of RBD-specific IgA, IgM and IgG kit. Testing results of RBD-specific IgA (A), IgM (B) and IgG (C) kit using 330 healthy sera, 138 interfering sera, 15 sera of once-suspected patients and 216 sera of 87 qPCR-confirmed COVID-19 patients. RLU: relative light units. Black bar indicates median values. The dotted line indicates the cut-off value. D-F: The receiver operating characteristic (ROC) curve analysis for SARS-CoV-2 diagnosis by the testing results of RBD-specific IgA, IgM or IgG (D, E and F, respectively) using 483 sera of SARS-CoV-2 negative individuals and 216 sera of SARS-CoV-2 infected patients. AUC, area under the curve of ROC.

Data from 216 sera samples were divided into 6 groups according to the time windows of collection after illness onset (**Table 2**). At 4-10 days after symptom onset, the RBD-specific IgA kit exhibited the highest positive diagnostic rate as 88.2% (15/17), while IgM and IgG kit showed detection rates of 76.4% (13/17) and 64.7% (11/17), respectively. The 2 sera diagnosed as negative in the 4-10 days group by IgA kit were collected at the 4th day after illness onset, which could be too early for detecting viral-specific antibodies of any types. In the group of 11-41 days after symptom onset, both RBD IgA and IgG kit showed the same positive diagnostic rate of 99.5% (198/199). In contrast, IgM kit somehow showed a relatively lower positive diagnostic rate as 98.5% (196/199). These results suggest that including IgA in a test provides better diagnostic outcome in early stage.

**Table 2.**
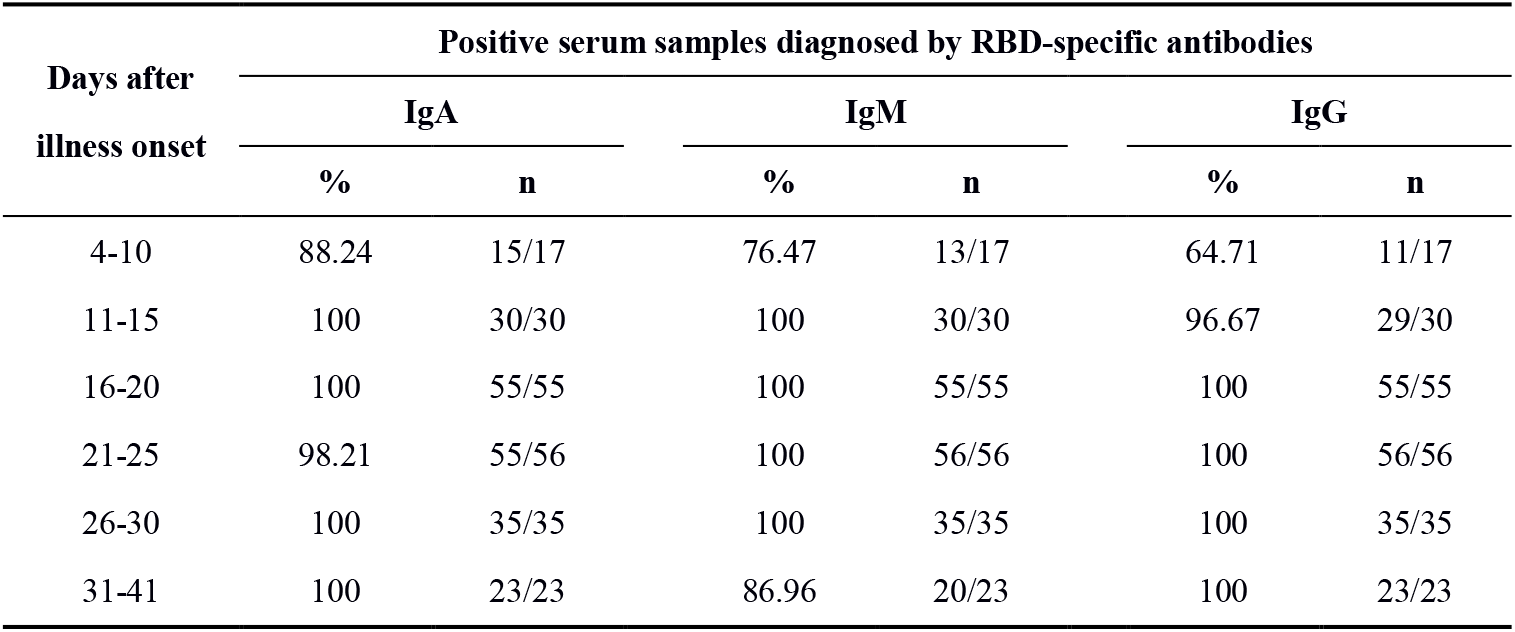
Sensitivity of RBD-specific IgA, IgM and IgG kits in serum samples obtained at different periods after illness onset.

| Days after<br>illness onset | Positive serum samples diagnosed by RBD-specific antibodies |  |  |  |  |  |
| --- | --- | --- | --- | --- | --- | --- |
|  | IgA |  | IgM |  | IgG |  |
|  | % | n | % | n | % | n |
| 4-10 | 88.24 | 15/17 | 76.47 | 13/17 | 64.71 | 11/17 |
| 11-15 | 100 | 30/30 | 100 | 30/30 | 96.67 | 29/30 |
| 16-20 | 100 | 55/55 | 100 | 55/55 | 100 | 55/55 |
| 21-25 | 98.21 | 55/56 | 100 | 56/56 | 100 | 56/56 |
| 26-30 | 100 | 35/35 | 100 | 35/35 | 100 | 35/35 |
| 31-41 | 100 | 23/23 | 86.96 | 20/23 | 100 | 23/23 |

### False positive analysis demonstrates the RBD-based detection are highly specific to SARS-CoV-2

When RBD-specific IgA, IgM or IgG individual kit was used, we observed a total of 9 (0.61% to 6.67%), 37 (5.54% to 40.0%) and 1 (0 to 0.73%) false positive cases in the three types of “negative controls”, respectively (**Table 3**). IgA kit was worse than IgG kit in yielding 9 false positive, but much better than IgM kit. Only one of the 9 cases who showed a weak positive signal (RLU was 38096) for IgA also tested weak positive (RLU was 22701) for IgM, other eight cases were tested negative for IgM and IgG kit. Except the only one case that was tested weak positive by IgA and IgM kit simultaneously, all other “negative controls” who had a positive signal for one isotype of antibody kit showed negativity for the rest of two other isotype antibody kit. Therefore, a combined test of using IgA and IgG (+/- IgM) kits can identify few false positive samples that show a positive signal for just one type of antibodies. The fact that very few cases of samples were IgA or IgG positive in 483 negative controls indicate that these RBD-based detection did not cross-interact with antibodies raised against other human coronaviruses (presenting in ∼15% of common cold cases and often causing pneumonia). Taken together, our RBD based CLIA kits are highly specific to SARS-CoV-2.

**Table 3.** Potentially false positive cases diagnosed by the current RBD-specific IgA, IgM and IgG kits.

| Antibody type | False-positive cases |  |  |  |  |  |
| --- | --- | --- | --- | --- | --- | --- |
|  | Healthy cases |  | Interfering cases |  | Once suspected cases |  |
|  | % | n | % | n | % | n |
| IgA | 0.61 | 2/330 | 4.35 | 6/138 | 6.67 | 1/15 |
| IgM | 5.45 | 18/330 | 9.42 | 13/138 | 40.0 | 6/15 |
| IgG | 0 | 0/330 | 0.73 | 1/138 | 0 | 0/15 |

For IgA detection, two persons of 330 sera collected previously from obviously healthy donors showed positive signals for no obvious reasons. For the interfering cases group who had various underlying diseases, four of six “positive” detected by the RBD IgA kits included two patients who had kidney disease and high-levels of self-antibodies, one had liver cirrhosis and another had breast cancer and received chemotherapy. The one “false positive” sample in the once suspected cases group was a breast cancer patient undergoing chemotherapy and had pneumonia.

### Kinetic studies of serum SARS-CoV2 RBD-specific IgA, IgM and IgG

We analyzed the kinetics of all the three antibody isotypes when multiple serum samples were collected from individual patients. Representative kinetic data from 9 COVID-19 patients was shown in **supplementary Figure 2**. To better understand the trends of antibody levels in all of the 87 COVID-19 patients (some of them contributed multiple samples), we plotted the median RLU reading according the time windows when sera were collected (**Figure 3A**), IgA detection show a highest sensitivity during about 4 to 25 days after illness onset. Because the RLU reading would vary among IgA, IgM and IgG due to different secondary antibodies used, we used highly purified RBD-specific IgA, IgM and IgG proteins from pooled sera of COVID-19 patients as standards (**supplementary Figure 3**). In this way, RLU reading was converted into absolute antibody concentration (amounts per mL). To simplify a plot from large numbers of samples, we only plotted median with interquartile range values of antibody concentrations vs. time windows. The median concentration of RBD-specific IgA reached the peak (8.8 μg/mL) during 16 to 20 days after illness onset, and then began to decline but remained at about 3.6 μg/mL until 31-41 days (**Figure 3B)**. The median concentration of RBD-specific IgG was the lowest in early disease stages but raised at 15 days post illness onset, the IgG concentration reached its peak during 21-25 days after illness onset as 16.5 μg/mL, and stayed at a relatively high concentration (11.4 μg/mL) until 31-41 days (**Figure 3B)**, suggesting that IgG is powerful for diagnostics at later stages. Although IgM concentration reached its peak (7.25 μg/mL) at early stages, it was lower than that of IgA or IgG. Our data suggest that IgM has the lowest diagnostic power among the three isotypes of antibodies for diagnosing SARS-CoV-2. Adding IgA into a diagnosis that contains IgG and IgM improves the serologic testing power at both early- and late-stage COVID-19.

**Figure 3.**
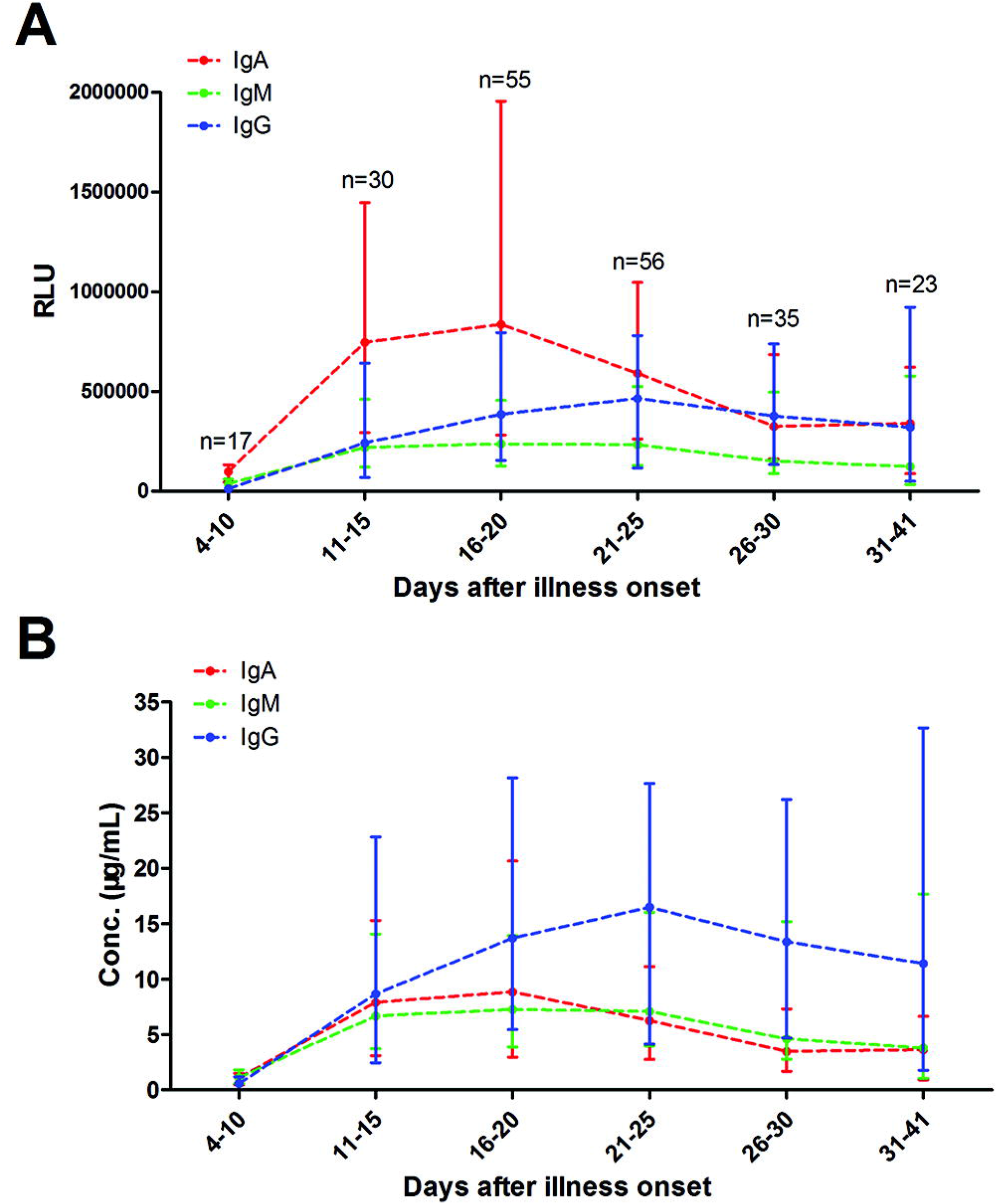
The kinetics of anti-RBD IgA, IgM and IgG levels in sera of COVID-19 patients at different time windows. The median values of RLU (A) or calculated antibody mass concentrations (B) were plotted for each isotypes of three antibodies, IgA (red), IgM (green) and IgG (blue). Bars indicate median with interquartile ranges.

### IgA as well as IgG and age positively correlated with COVID-19 severity

To explore whether a simple laboratory test such as measuring RBD-specific antibody levels in serum could serve as a quantifiable indicator for COVID-19 severity, we divided the 87 patients into three severity groups based on established clinical classifications. Consistent with previous studies [29], we found that disease severity was positively correlated with age in our cohort (**supplementary Figure 4**). Patients with severe symptoms were significantly older (median age of 62.5 years) than those patients with moderate (median age of 46 years) and mild symptoms (median age of 30 years). Remarkably, we found that IgA concentrations in severe cases were significantly higher than those mild or moderate cases (**Figure 4A**). IgG levels in moderate and severe COVID-19 patients were also higher than mild cases (**Figure 4C**), which was previously reported [9, 20].

**Figure 4.**
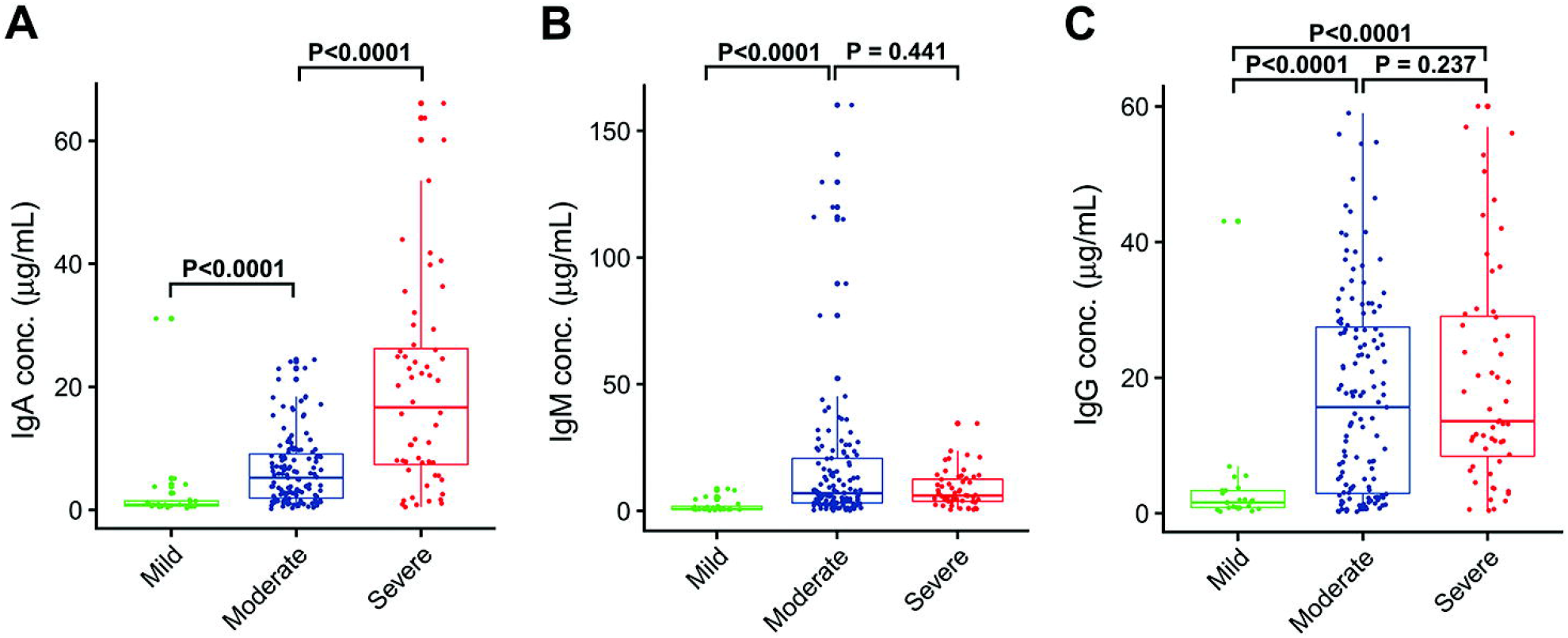
Serum antibody levels in three distinct severity groups of COVID-19 patients. Mild: 25 sera from 9 patients; moderate: 135 sera from 56 patients; and severe: 56 sera from 22 patients. 5 critical patients were merged into the severe group. Antibody levels in serum samples were collected from confirmed patients at 4 - 41 days post illness onset and presented as scatter plots. For IgA (A), levels in mild, moderate and severe patients were sequentially increased (p values indicated). Results for IgM are shown in B. For IgG (C), levels in moderate and severe patients were significantly higher than mild patients.

## Discussion

The nucleocapsid protein (NP) is the most abundant protein in coronaviruses, and often used as a diagnostic marker for coronaviruses such as SARS-CoV[30]. The RBD is the ligand for receptor ACE2, therefore RBD could be a main target for neutralization antibodies[31, 32]. Here, we explored the possibility of using either NP or RBD as an immobilized antigen for developing a clinical COVID-19 diagnostic test. Our data (**Figure 1**) demonstrated RBD-based test is better than NP-based test. One possible mechanisms of difference is that the NP antigen is expressed in bacteria, whereas the RBD protein used here is expressed in a human cell line enabling critical glycosylation for high-affinity binding to antibodies produced in COVID-19 patients.

When we combined our RBD-specific IgA and IgG kits together, the sensitivity, specificity and overall agreement elevate to 99.1%, 100% and 99.7%, respectively (**Table 1**). Thus, our serological tests have much higher accuracy than the current methods of detecting viral RNA (sensitivity < 70%) [8-12], and published immune-assays such as “flow immunoassay” and ELISA in earlier studies [9-11, 16-20, 22, 23]. We revealed that both IgM and IgA had early responses, while IgG showed up later. Rapid increase of the three isotypes of serum RBD-specific antibodies started at about 10 days after illness onset (**supplementary Figure 2A-C**). The early appearance of IgA in COVID-19 patients’ sera is probably due to the initial infection of this virus at the respiratory system enriched with mucosal immune cells. The low basal level of IgA in serum makes SARS-CoV-2 specific IgA detection highly sensitive at early stage of infection. Therefore, IgA should be included in a serological test, which may provide higher diagnostic accuracy for COVID-19.

When we analyzed IgA, IgM or IgG concentrations in the sera of patients with different severity, we observed that disease severity was positively correlated with serum IgA concentrations **(Figure 4A**). The underlying mechanisms of this novel observation need to be further investigated in the future. IgA is traditionally recognized to play an anti-inflammatory role and prevent tissue damage at mucosal sites. However, recent reports also demonstrated that serum IgA is involved in the formation of immune complexes to amplify inflammatory responses[33]. Serum IgA induced proinflammatory cytokine production by macrophages, monocytes and Kupffer cells in non-mucosal tissues including liver, skin and peripheral blood[34]. The latter phenomenon indicates possible antibody-dependent enhancement (ADE) of infection. The immunopathological effects of ADE have been observed in various viral infections, characterized as antibody-mediated enhancement of viral entry and induction of a severe inflammatory response[35]. It is unclear currently whether IgA as well as IgG contributed directly (e.g. via ADE) or indirectly (e.g. leading to a pathogenic inflammatory storm[36]) to the worse clinical outcome in severe COVID-19 patients. If so, blocking of IgA-Fc alpha Receptor I (FcαRI, CD89, an IgA receptor) interaction could mitigate ADE or inflammatory storms, thus providing a novel treatment strategy.

The current study has several limitations. Serum samples were not available every day for each patient, the earliest serum was collected at the 4th day, and last one was at the 41th day after self-reported illness onset. There are only 17 cases of serum samples collected within the first 10 days after illness onset; which consequently influenced the accuracy of early. Similarly, there were only 23 cases of serum samples taken after 30 days post illness onset, hampering an analysis of long-term antibody levels in recovered patients. Most patients enrolled in this study were with clinically moderate symptoms (56/87, 64.4%). There were 17 severe and 5 critical cases, respectively. There were also few cases of mild COVID-19 patients. Therefore, this study of the correlation between antibody levels and disease severity warrants further investigation.

In summary, this study reports a novel serological test for detecting SARS-CoV-2 RBD-specific IgA as well as IgM and IgG for clinical diagnosis of COVID-19. Due to its high specificity and sensitivity, this approach could sensitively and quantitatively measure levels of the three types of antibody in blood and other tissues. The serological study also provides valuable information for monitoring SARS-CoV-2 infection, understanding of COVID-19 pathogenesis and improving strategies for preventing, treating and vaccine development of this pandemic disease.

## Data Availability

All data referred to our manuscript is available.

## Acknowledgements

We would like to thank the staff and patients at Department of Infectious Diseases, The First Affiliated Hospital of USTC for their support in providing samples and clinical data collection. We would also like to thank Profs. Jianping Weng and Tian Xue and other colleagues in Division of Life Sciences and Medicine for their generous and professional support. We would like to thank Prof. Yan Xiang at University of Texas Health Science Center at San Antonio for critical reading and comments on this manuscript. We would specially thank Prof. Peihui Wang at Shandong University for a plasmid expressing the SARS-CoV-2 spike protein.

## Authorship Contributions

Tengchuan Jin, Yajuan Li and Xiaoling Ma provide funding, designed the study, participated in data analysis, and wrote the manuscript. Huan Ma, Weihong Zeng and Hongliang He designed the study, performed the majority of experiments, analyzed the data and drafted the manuscript. Other authors participated in the experiments and/or writing of the manuscript.

